# Defining a subgroup within autism leads to potential enrichment criteria in investigational therapeutic development: Comparing resting state gamma band power to a fragile X syndrome reference sample

**DOI:** 10.64898/2026.10.06.26364877

**Authors:** Grace C. Westerkamp, Stella T. Sarraf, Peter Vanderklish, Sharron Gargosky, Rana Elmaghraby, Ernest V. Pedapati, Craig A. Erickson

## Abstract

**Background:** Elevated resting gamma power is a reproducible, robust neurophysiology marker across species in fragile X syndrome (FXS), a major known cause of autism spectrum disorders (ASD). In FXS, this neurophysiological phenotype is well described and particularly prominent in males harboring fully methylated full CGG repeat expansions [1,4,16–20]. However, gamma power evaluations in idiopathic ASD have yielded variable results. As a single-gene model of ASD, FXS hold promise to biologically model some of the autism spectrum. Exaggerated gamma power in FXS is thought to reflect increased cortical excitability, a common feature of ASD, and has been shown to correlate with symptoms in both human FXS individuals and the Fmr1 knockout mouse model of FXS [1,19,20]. A recent clinical trial of the BK channel positive modulator, Codabakalner, provided encouraging signs that normalization of gamma power in FXS may provide therapeutic benefit [14]. Interventional and biomarker studies in ASD provide support that this may also be the case in ASD [15]. We utilized gamma band power to evaluate neurophysiological overlap between ASD and FXS seeking to determine the prevalence and clinical characteristics of those with ASD who exhibit a FXS-like elevated resting state gamma phenotype.

**Methods:** This cross-sectional resting EEG analysis included 159 individuals with non-syndromic ASD and 41 males with fully methylated full mutation FXS. Relative gamma power was calculated as 30– 100 Hz power divided by 1–100 Hz power across 10–20-equivalent electrodes, excluding 55–65 Hz. We defined the cutoff for elevated resting gamma band power in ASD as relative gamma power of greater than or equal to the median minus one standard deviation of gamma power in our FXS male sample.

**Results:** A FXS-derived relative-gamma threshold of 0.185 or greater was met by 22% of participants with ASD (n=35). As expected, the mean ASD group resting state gamma was lower than the male FXS sample (p < 0.001). ASD participant threshold proportions by age group were 3/37 (8.1%) below 13 years, 9/50 (18.0%) age 13-18 years, and 23/72 (31.9%) in the 18+ years group. Within the ASD group, there were no observable clinical characteristics that readily differentiated participants with an elevated gamma phenotype from those without this phenotype.

**Conclusions:** Approximately one fifth of all participants and slightly over 30 percent of adults with ASD met the FXS-derived elevated resting gamma band power criterion. This represents the first investigation using a translational brain neurophysiology marker in a syndromic form of autism to parse a clinical idiopathic ASD sample to identify potential overlapping brain features. The results will work to support potential targeting of ASD subgroups in the future utilizing therapeutic approaches initially established in a single-gene form of developmental disability.

## Background

Resting-state electroencephalography (EEG) provides a way to characterize neural activity across neurodevelopmental conditions without requiring a complex task. In participants with fragile X syndrome (FXS), Wang and colleagues reported elevated resting state gamma band power and an association with social-communication difficulties [1]. Here, we investigated a group of adolescent and adult male participants with FXS, a population repeatedly demonstrated to have elevated resting state gamma band power, and compared them to a heterogenous group of individuals with idiopathic autism to evaluate the potential prevalence of gamma band power elevation in non-syndromic autism spectrum disorder (ASD). This comparison is the first effort to evaluate neurophysiologic pathology that may be shared between a characteristic group of individuals with a single gene cause of developmental disability (FXS) and a heterogenous group with ASD.

Literature describing high-frequency gamma band activity in individuals with ASD is mixed. Cornew and colleagues reported regional elevations in high-frequency gamma power in children with ASD using eyes-closed magnetoencephalography [2]. Additional EEG studies have reported increased resting-state gamma power and atypical gamma laterality in ASD [3,21,22]. In contrast, Maxwell and colleagues found lower resting-state EEG gamma band power at right lateral electrodes in boys with ASD [3]. Differences in modality, spatial sampling, and resting conditions limit direct comparison of these findings. They also motivate an individual-level description alongside group-level comparisons, rather than assuming that all participants with ASD share the same gamma band pattern.

Demographic and measurement characteristics further constrain interpretation. Relative EEG power in FXS varies by sex in several frequency bands, including gamma [4]. In addition, a conventional frequency-band measure combines contributions from oscillatory and non-oscillatory (aperiodic) spectral activity unless these components are explicitly separated [5]. Relative gamma should therefore be described according to its measured definition, without equating a high ratio with a specific cellular mechanism.

The present study asked what proportion of an ASD sample met a relative-gamma criterion derived from an all-male FXS reference group, and how that proportion varied across age strata and clinical presentation. The reference criterion was the FXS median resting relative gamma band power minus one sample standard deviation, applied as a lower bound without an upper limit. This approach was intended to characterize overlap with the selected FXS reference group while estimating the prevalence of an FXS-like biological subtype in the wider population with ASD.

Within the ASD group, we additionally examined whether participants with elevated gamma band power presented with an observable phenotypic difference than those without this EEG feature.

## Methods

### Study design and setting

We conducted a cross-sectional analysis of available eligible resting-state EEG recordings collected at our lab during a ten-year period from August 2015 through July 2025.

### Participants and recording selection

The final cohort comprised 159 participants with ASD and 41 unique fully methylated full mutation male participants with FXS. For ASD, each participant’s most recent valid resting-state EEG was selected, defined as a recording that had not been rejected because of heavy artifact contamination. Availability of clinical scores did not determine inclusion in this EEG analysis. The ASD group included both sexes and all ages represented in the selected cohort in order to represent a general, heterogenous autism population. In all cases, the diagnosis of ASD was made utilizing DSM V criteria and use of the Autism Diagnostic Observation Schedule (ADOS).

All participants with FXS had full-mutation full methylated FXS confirmed by Southern blot analysis and polymerase chain reaction. FXS inclusion required a valid resting-state EEG and confirmed full-mutation status. The FXS reference group comprised males aged at least 13 years for subjects with multiple EEGs on file, the most recent valid EEG data was utilized. Each of the 41 participants with FXS contributed one recording. EEG recordings rejected for heavy artifact contamination were excluded.

### EEG acquisition and preprocessing

The EEG acquisition protocol specified five minutes of EEG while participants watched a silent video, using an EGI NetAmp 400 amplifier and 32- or 128-channel HydroCel electrode nets at 1000 Hz. Preprocessing used AutoClean EEG Pipeline version 3.0.0 and included resampling to 250 Hz, filtering at 0.5–100 Hz, line-noise filtering at 55–65 Hz, average referencing, bad-channel identification and spherical-spline interpolation, and movement-artifact rejection. Extended-infomax independent component analysis was used to identify and remove components attributed to ocular, muscle, channel, and cardiac artifacts based on their temporal, spatial, and spectral features. Data were segmented into two-second epochs and manually reviewed for quality.

Retained epoch counts were summarized for each selected recording. Analyses used 20 channels mapped to 10–20 electrode positions for both net types. The net-specific electrode mappings are provided in Additional file 1, Table S4.

### Relative gamma power

Power spectral density (PSD) was estimated using Welch’s method in MNE-Python with a nominal two-second segment length, a one-second overlap setting, and the default taper. Spectra were averaged arithmetically across retained epochs and the 20 mapped channels before calculating the band-power ratio. Channel selection occurred after average referencing; it did not constitute rereferencing to a 10–20 montage.

Relative gamma was calculated as the sum of power from 30 through 100 Hz divided by the sum from 1 through 100 Hz, omitting the inclusive interval 55–65 Hz from both sums. Thus, 100 Hz was included and 65 Hz was excluded. The resulting unitless ratio describes the fraction of retained broadband power in the gamma range. No periodic–aperiodic separation was applied.

### ASD Elevated Gamma Threshold Classification

For the present ASD classification, we used a distinct threshold: the median relative gamma of the selected FXS recordings minus one sample standard deviation (denominator n − 1). Values greater than or equal to this threshold were classified as meeting the reference criterion, with no upper bound. Relative-gamma values are displayed to three decimal places; calculations and threshold classification used full precision. The term “FXS-comparable” denotes only this operational threshold; it does not imply statistical equivalence, shared etiology, or clinical similarity. The threshold was neither optimized nor independently validated as a diagnostic classifier.

### Clinical assessments

Within the ASD group, we compared the clinical phenotype of participants with and without the elevated resting state EEG gamma feature. For this comparison, 35 participants with ASD met the elevated gamma band power criteria and 124 did not. Missing clinical measure scores were omitted separately for each measure without imputation; clinical data availability was not required for inclusion in the EEG analysis ASD cohorts.

Six assessment sets contributed 27 measures. Cognitive outcomes included the overall IQ composite (standard score), available IQ estimate (standard score; hierarchical), verbal-domain standard score, and nonverbal-domain standard score when available. When multiple IQ data points were available per participant, we prioritize the overall composite IQ when available followed by abbreviated IQ and then non-verbal IQ for those unable to generate verbal or full-scale IQ scores. In all, 123 composite IQ, 5 abbreviated IQ values, and one non-verbal domain score only were included.

Other clinical measures used in our ASD group comprised of: seven Social Responsiveness Scale (SRS) raw score domains, Social Communication and Interaction (SCI), Social Awareness, Social Cognition, Social Communication, Social Motivation, and Restricted/Repetitive Behavior [7]; five Aberrant Behavior Checklist (ABC) raw subscale scores (Irritability, Lethargy/Social Withdrawal, Stereotypy, Hyperactivity, and Inappropriate Speech) [8]; the Social Communication Questionnaire (SCQ) Lifetime raw total [9]; and four Vineland Adaptive Behavior Scales standard scores (Adaptive Behavior Composite, Communication, Daily Living Skills, and Socialization) [10].

### Statistical analysis

We summarized age using the mean, standard deviation, median, and range, and relative gamma using the median and interquartile range (IQR). Threshold membership was summarized as a count and percentage. Two-sided asymptotic Mann–Whitney U tests with continuity correction compared continuous distributions. The standalone ASD–FXS comparisons for the full group and the ASD group aged at least 13 years were unadjusted. Benjamini–Hochberg false-discovery-rate correction was applied separately to the two pairwise comparisons among ASD and FXS; the three ASD-age-stratum–FXS comparisons; and the two pairwise group comparisons within each sensitivity panel.

ASD age strata were below 13 years, 13-18 years, and 18 years or older. The ordered trend in continuous relative gamma was tested using the two-sided normal approximation to the Jonckheere–Terpstra statistic, without continuity correction; no outcome ties were present. This test evaluated relative-gamma distributions across ordered strata, not threshold-membership proportions.

Sensitivity analyses restricted all groups to males, age at least 13 years, males aged at least 13 years, and males aged at least 18 years. Each panel recalculated the threshold from its eligible FXS reference group. Age restriction did not equate the groups’ age distributions. Analyses did not adjust for continuous age, medications, cognitive ability, study, recording net, or family relatedness. No longitudinal analyses were included. The analysis plan was specified before analyses were run and results reviewed; it was not formally registered.

Clinical scores were summarized by group (elevated or non-elevated gamma ASD) using the mean and standard deviation with measure-specific sample sizes.

## Results

### Group characteristics

The analysis included 200 unique participants: 159 with ASD (males and females) and 41 in the FXS (all male) reference group (**Table 1**). The ASD group included 142 males and 17 females and had a mean age of 16.90 years (SD 5.76) and the FXS group had a mean age of 29.66 years (SD 9.15). The groups therefore differed substantially in age and sex composition.

**Table 1.** Participant characteristics at the selected EEG.

| Group | n | Male/female, n | Age, mean (SD) | Median age | Age range |
| --- | --- | --- | --- | --- | --- |
| ASD | 159 | 142/17 | 16.90 (5.76) | 17.08 | 0.59–39.83 |
| FXS reference | 41 | 41/0 | 29.66 (9.15) | 27.73 | 13.77–50.33 |
| ASD with elevated gamma power | 35 | 33/2 | 18.86 (5.39) | 19.85 | 5.19–31.96 |

Ages are in years. The ASD subgroup is contained within the full ASD cohort. The FXS reference includes 41 unique participants; related individuals remain in the cohort.

Retained resting-state EEG epoch counts were available for all 275 selected recordings. Median counts were 124 (IQR 116–140; range 78–375) for ASD and 112 (IQR 97–169; range 31–245) for the FXS reference group.

### Relative gamma and reference-threshold membership

The FXS median relative gamma was 0.254 and the sample standard deviation was 0.070, yielding a threshold for elevated gamma power of 0.185. Thirty-five of 159 participants with ASD (22.0%) met this criterion (**Table 2**; **Figure 1**). This subgroup included 33 males and two females and had a mean age of 18.86 years (SD 5.39).

**Figure 1.**
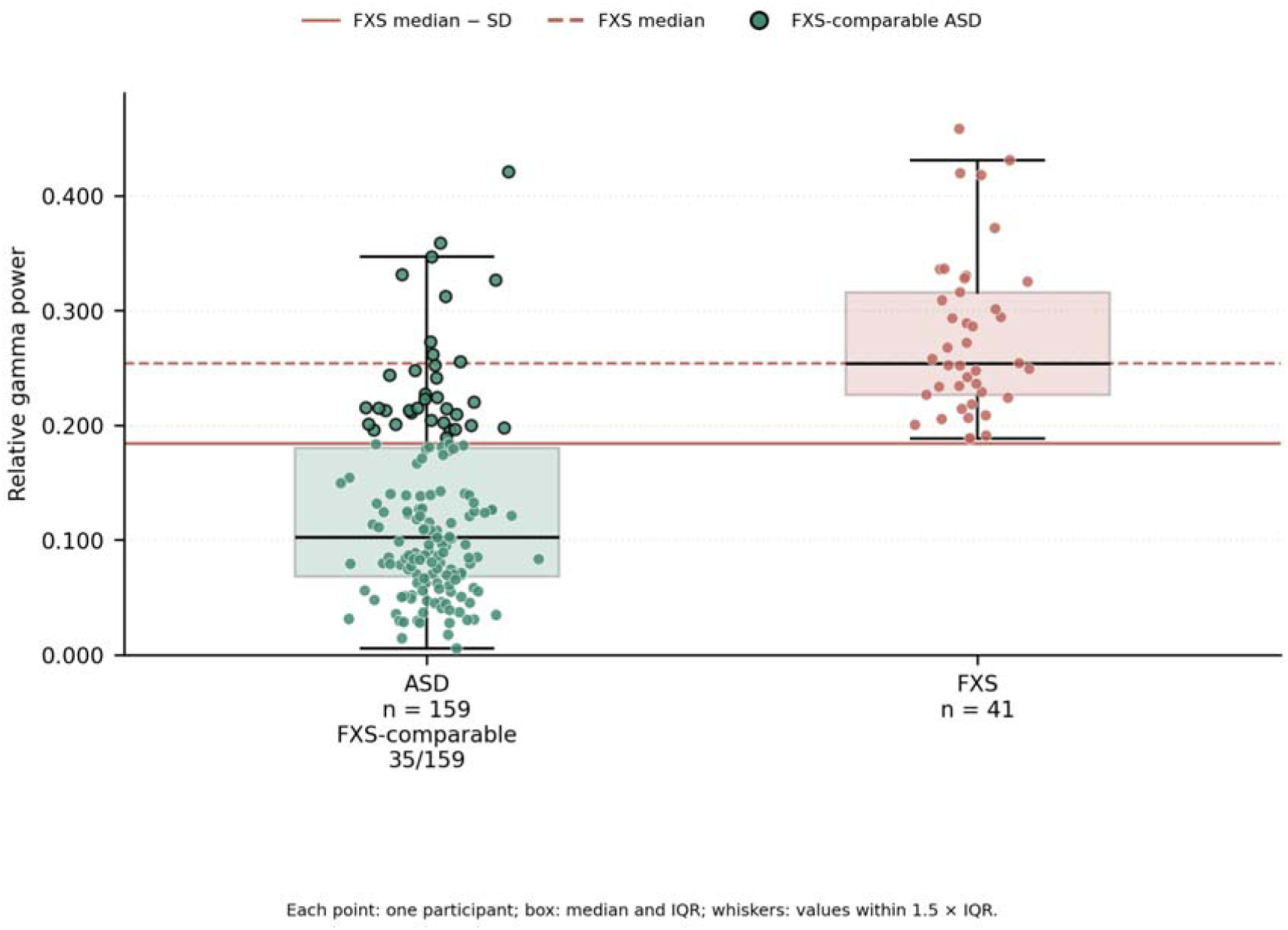
Relative gamma in ASD and the FXS reference group. Relative gamma from one selected EEG per participant is shown for ASD (n = 159) and the FXS reference group (n = 41). The FXS-comparable ASD subgroup comprised 35/159 participants (22.0%) meeting the FXS median-minus-SD criterion (0.185). FXS-comparable denotes meeting the EEG reference criterion, not clinical equivalence; black-outlined ASD points identify these participants. The FXS median was 0.254. Points, boxes, and reference-line keys are defined in the graphic. The two-sided Mann–Whitney comparison yielded U = 472, p < 0.001 (unadjusted). The FXS cohort and its selected recordings were conditioned on elevated relative gamma.

**Table 2.** Relative-gamma distributions.

| Group | n | Median | 25th–75th percentile |
| --- | --- | --- | --- |
| ASD, all subjects | 159 | 0.103 | 0.068–0.180 |
| FXS | 41 | 0.254 | 0.227–0.316 |

Values are unitless fractions. The FXS reference group was selected for the known elevated relative gamma in males with this gene cause of autism. The full-sample ASD–FXS gamma comparison test gave U = 472, p < 0.001 (unadjusted).

### Age-restricted and age-stratified findings

Among 122 participants with ASD aged at least 13 years, 32 (26.2%) met the elevated gamma power reference criterion (**Figure 2**). The ASD–FXS distribution comparison remained significant (U = 409, p < 0.001). The age-restricted ASD group had a mean age of 19.17 years (SD 4.33), compared with 29.66 years in FXS.

**Figure 2.**
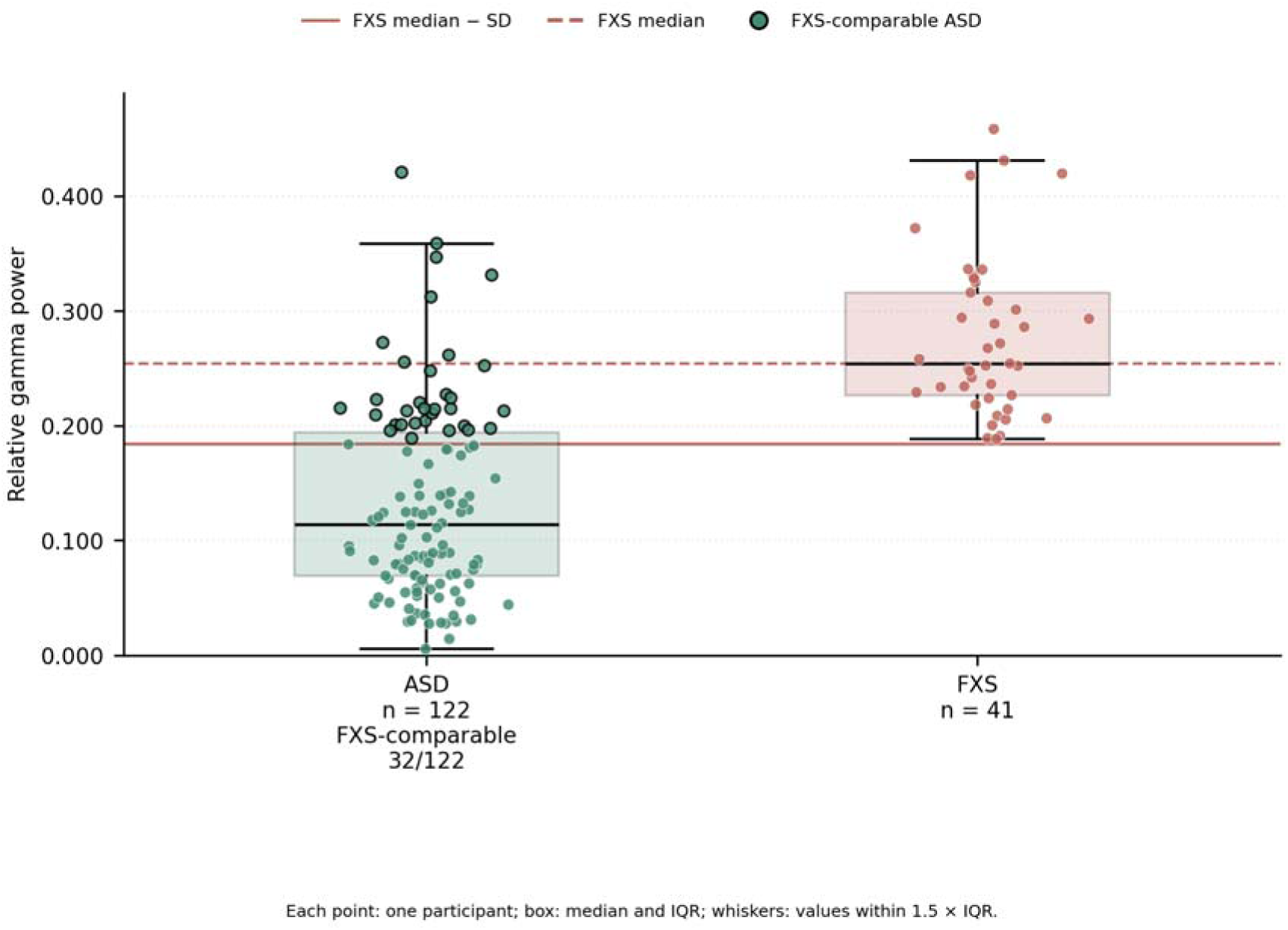
Relative gamma after restriction to age 13 years or older. The ASD group aged at least 13 years (n = 122) is compared with the FXS reference group (n = 41). The FXS-comparable ASD subgroup comprised 32/122 participants (26.2%) meeting the full-reference criterion of 0.185. FXS-comparable denotes meeting the EEG reference criterion, not clinical equivalence; black-outlined ASD points identify these participants. The two-sided Mann–Whitney comparison yielded U = 409, p < 0.001 (unadjusted). The restriction did not match the age distributions of the two groups.

The proportions meeting the criterion were 3/37 (8.1%) below age 13, 9/50 (18.0%) at 13 to under 18, and 23/72 (31.9%) at age 18 or older (**Table 3**; **Figure 3**). Continuous relative gamma showed an ordered trend across these strata (Jonckheere–Terpstra J = 5120, z = 3.412, two-sided p = 0.000646). Each ASD stratum differed from the selected FXS distribution after adjustment (all adjusted p < 0.001). The stratum-specific proportions are descriptive; the trend test concerned the continuous outcome.

**Figure 3.**
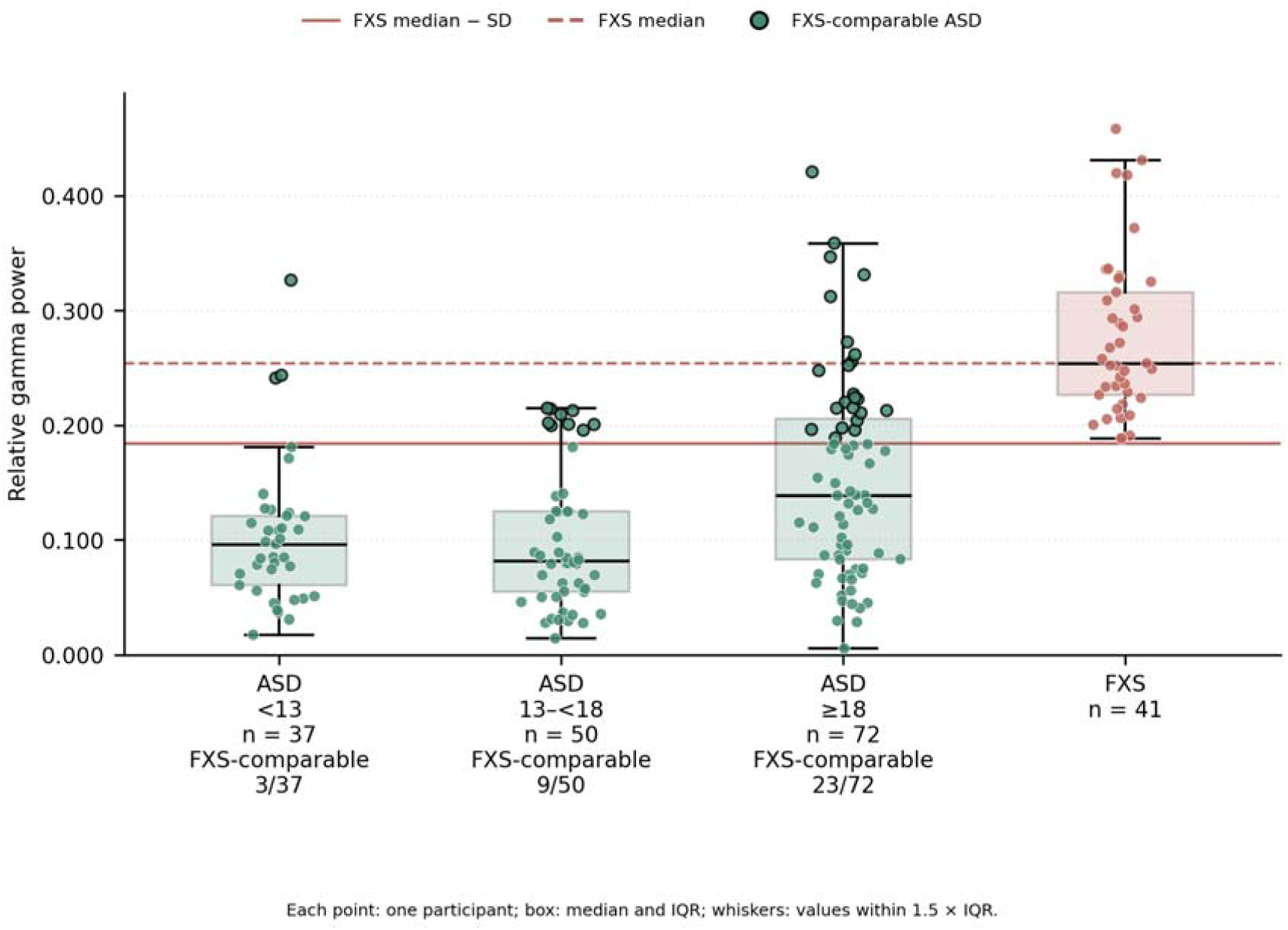
Relative gamma across ASD age strata. Participants with ASD are shown below age 13 (n = 37), at 13 to under 18 (n = 50), and at age 18 or older (n = 72), alongside the FXS reference group (n = 41). The same full-reference threshold was applied to every ASD stratum, identifying FXS-comparable subgroups of 3/37 (8.1%), 9/50 (18.0%), and 23/72 (31.9%) participants, respectively. FXS-comparable denotes meeting the EEG reference criterion, not clinical equivalence; black-outlined ASD points identify these participants. The two-sided Jonckheere–Terpstra test of continuous relative gamma gave J = 5120, z = 3.412, p = 0.000646. Each stratum differed from FXS after correction across the three comparisons (all adjusted p < 0.001). These are cross-sectional comparisons, and the trend test does not test the threshold-membership proportions.

**Table 3.**
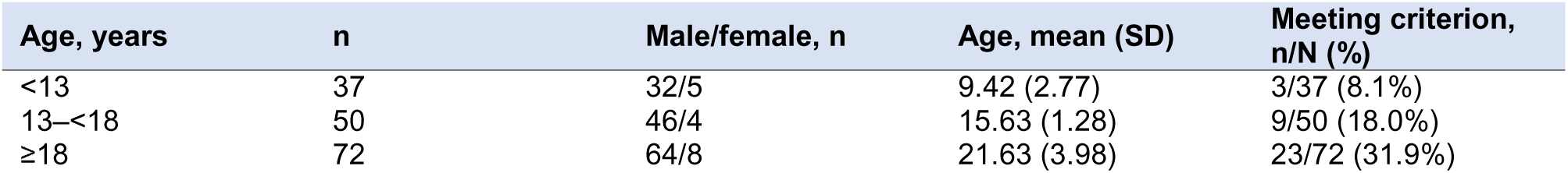
ASD age strata and reference-threshold membership.

The same threshold (0.185) was applied across strata. Membership proportions are descriptive. The Jonckheere–Terpstra test concerns continuous gamma, not these proportions.

### ASD subjects meeting gamma threshold by sex and age

The proportions of participants with ASD meeting the panel-specific criterion were 33/142 (23.2%) among males, 32/122 (26.2%) among participants aged at least 13 years, 30/110 (27.3%) among males aged at least 13 years, and 21/64 (32.8%) among males aged at least 18 years (**Additional file 1, Table S1 and Figure S2**). The first three panels retained the full FXS threshold of 0.185.

### Clinical assessment findings within ASD

Clinical assessment availability differed between participants and measures; 35 participants with ASD met and the 124 did not meet the relative-gamma reference criterion (**Table 4**).

**Table 4.** Clinical assessment availability in the ASD group (available/total number participants)

| Assessment | Elevated gamma group | Below gamma criterion | Total with data |
| --- | --- | --- | --- |
| IQ | 29/35 | 100/124 | 129 |
| SRS | 29/35 | 89/124 | 118 |
| ABC | 30/35 | 101/124 | 131 |
| SCQ Lifetime | 23/35 | 90/124 | 113 |
| Vineland | 27/35 | 84/124 | 111 |

Meeting criterion denotes relative gamma at or above the reference threshold (displayed as 0.185); classification used the full-precision threshold. IQ includes several test families and chart sources. SRS and ABC use raw scores, SCQ uses Lifetime total score, and the Vineland uses four standard scores. Complete score-specific results appear in (**Additional file 1, Tables S3– S7**).

None of the 27 clinical-score comparisons between the two ASD groups survived assessment-specific correction (**Additional file 1, Table S3**). Complete results for all six assessments, including coefficients, group summaries, sample sizes, and unadjusted and corrected p-values, are provided in the supplement. (**Tables S6–S7**).

## Discussion

FXS-informed resting state EEG may help characterize neurophysiological heterogeneity within ASD. Elevated resting relative gamma power distinguished a subgroups of participants with ASD who shared this electrophysiologic feature with fully methylated full mutation FXS males. The greater representation of this phenotype was noted in the older age strata which further highlights age as an important consideration interpreting this overlap. Overall, we demonstrated that a subgroup of individuals with ASD exhibit an elevated resting EEG gamma phenotype that aligns with the resting EEG pathophysiology well established in fully methylated full mutation adolescent and adult males with FXS.

The use of an FXS reference group was motivated by prior reports of elevated resting gamma in FXS [1]. Previous ASD studies have reported both regional high-frequency increases in resting magnetoencephalography and regional decreases in resting EEG gamma [2,3]. The present analysis adds an individual-level threshold description to a group-comparison framework.

The FXS reference group deliberately only included fully methylated full mutation males given this population has consistently demonstrate elevated resting gamma power. This finding, which is also noted in the FMR1 KO mouse, is not as consistent nor as clear in repeat expansion size mosaic males and in females in this X linked single-gene cause of autism thus supporting a molecular dosing effect on gamma pathophysiology in FXS. This approach limits phenotypic heterogeneity within a single gene model of ASD, thus strengthening our ability to parse ASD using a comparison group enriched for elevated gamma band activity. Consequently, the large separation between ASD and this reference should not be interpreted as an unbiased comparison between ASD and the entire FXS population. Moreover, the observed ASD fraction depends on the definition of the reference, the selected session, and the median-minus-standard-deviation approach.

The older ASD subject strata had larger descriptive proportions above the gamma threshold. The continuous-outcome trend supports a cross-sectional association with age stratum, but it cannot establish that relative gamma increases within individuals over development. Cohort composition and other age-associated characteristics could contribute. Restricting ASD to age at least 13 years retained the threshold-defined subgroup pattern while leaving the ASD group substantially younger than FXS.

The limited representation of females in ASD and their absence from both comparison groups constrain generalization across sexes. Prior evidence of sex-related differences in FXS relative EEG power reinforces the need to evaluate sex explicitly in future work [4].

Relative gamma expresses gamma-band power as a fraction of broadband power. A higher ratio can arise from differences in gamma power, power elsewhere in the denominator, or both. Because the analysis did not separate periodic and aperiodic components, it cannot attribute the findings specifically to stronger gamma oscillations versus aperiodic activity [5]. In males with FXS, we and others have established that increased relative gamma power does co-occur with cortical excitability and increased E/I balance as measured by fitting oscillations and one over F (FOOOF) analysis. Future analysis within the elevated gamma power ASD subgroup should include evaluations of all power bands and periodic/aperiodic FOOOF analyses. Scalp high-frequency EEG measurements are also vulnerable to muscle contamination; experiments using neuromuscular blockade provide direct evidence of this concern [12]. Artifact rejection, independent component removal, and manual review were included in preprocessing, and retained epoch counts are reported for each diagnostic group.

Regarding the clinical assessments in the ASD group, across the six assessment sets, the analyses did not identify differences between the two ASD groups that survived assessment-specific correction. While these results do not establish that the groups are clinically equivalent, there are no readily observable clinical differences between the high and low gamma ASD subgroups. The gamma biologic marker may be identifying a subgroup within ASD that is not readily externally observable based on clinical severity or general clinical presentation. This points to how current behavioral outcomes may be insufficient to parse biological heterogeneity in ASD and this will likely have and important impact on future therapeutics development in autism. This is also in alignment with the many large-scale controlled drug trial in autism in the last 25 years that have largely included participants based on observable clinical criteria only. The EEG overlap motivates testing whether interventions that modify gamma-related physiology in FXS could also benefit participants with ASD meeting the relative-gamma criterion. In acute placebo-controlled crossover studies in FXS, baclofen reduced resting gamma in 17 participants [13], and codabakalner reduced 30–50 Hz gamma in 10 adult men, alongside significant improvement on the NIH Toolbox Flanker task [14]. These findings suggest potential gamma target engagement treatment-responsive physiology in FXS. The present analysis does not specifically establish a shared mechanism between an elevated gamma ASD subgroup and fully methylated full mutation males with FXS though this does point towards the potential for shared cortical hyperexcitability in these participant groups. Future prospective placebo-controlled studies could compare therapeutics effects in males with FXS and a sub-group of participants with non-syndromic ASD selected for participation based on baseline gamma resting EEG findings. In fact, the next trial of codabakalner in neurodevelopmental disorders (NCT07439510) plans to enroll adult full mutation fully methylated males with FXS and a separate cohort of participants with non-syndromic (idiopathic) ASD who are found during screening EEG to exhibit the elevated resting EEG gamma phenotype that characterizes males with FXS and is described in this paper. To our knowledge this clinical trial application utilizing the overlap of elevated gamma band power across FXS and ASD to parse an autism participant population is the first prospective use of neurophysiology inclusion criteria in neurodevelopmental disorders therapeutics development. This methodology builds upon the published work describing the impact of codabakalner use in FXS to hypothesize what ASD sub-population may best potentially respond to this novel treatment approach.

### Limitations

This subject EEG group was assembled from existing research recordings, so the proportion meeting the criterion should not specifically be interpreted as well-defined population prevalence. Additionally, cross-sectional age patterns cannot establish developmental change within individuals.

Female participants were underrepresented: 17 of 159 participants with ASD were female, and the FXS and control groups were entirely male. Greater female representation is needed to assess whether these findings extend across sexes. The groups also differed substantially in age, and analyses did not adjust for age, medication, cognitive ability, study, or recording net. Related individuals remained in the groups, and analyses did not account for family dependence.

Clinical data were incomplete and varied in instrument, respondent, assessment timing, and documentation. Available-case analyses therefore represented different subsets of the ASD EEG cohort across measures.

## Conclusions

Elevated resting relative gamma characterized a subgroup of participants with ASD, suggesting electrophysiological overlap with the selected male FXS reference group.The presence of elevated gamma power was descriptively more frequent in older ASD strata. Within ASD, no threshold-group differences were noted in the elevated gamma subgroup compared to others with idiopathic ASD. This analysis marks an initial step in efforts to evaluate neurobiological overlap between syndromic and idiopathic forms of autism to aid future therapeutic planning helping to parse heterogeneity in ASD that has confounded treatment development for decades.

## Data Availability

All data produced in the present study are available upon reasonable request to the authors if agreed to by the sponsor this research analysis.

## List of abbreviations

ABC: Aberrant Behavior Checklist
ADAMS: Anxiety, Depression, and Mood Scale
ADOS: Autism Diagnostic Observation Schedule
ASD: autism spectrum disorder
DSM: Diagnostic and Statistical Manual of Mental Disorders
EEG: electroencephalography
FDR: false discovery rate
FXS: fragile X syndrome
IQ: intelligence quotient
IQR: interquartile range
PSD: power spectral density
SCI: Social Communication and Interaction
SCQ: Social Communication Questionnaire
SD: standard deviation
SRS: Social Responsiveness Scale.

## Declarations

### Ethics approval and consent to participate

The Institutional Review Board at Cincinnati Children’s Hospital Medical Center approved the research procedures. The study protocols specified written informed consent from participants able to provide consent or from a parent or legally authorized representative, with participant assent when appropriate and feasible.

### Consent for publication

Not applicable.

### Availability of data and materials

Study data and analysis code are available from the corresponding author, Grace C. Westerkamp, upon request and in consultation with the study sponsor Spinogenix, Inc.

## Authors’ contributions

GCW contributed to EEG data collection, performed EEG preprocessing and data analysis, and drafted the manuscript. CAE, RE, and EVP: Writing – review & editing and conceptualization of the primary methods, hypotheses, and data use planning. SS, SG, and PK participated in the conceptualization of the project and reviewed and edited the manuscript.

## Acknowledgements

We thank the participants who took part in the research studies at Cincinnati Children’s Hospital Medical Center that contributed to this analysis.

## Supplementary Material

### Additional file 1: supplementary EEG and clinical analyses

Companion to: An EEG-defined relative-gamma subgroup in autism spectrum disorder: frequency and clinical characterization using a selected fragile X syndrome reference

Table S1. **Comparisons within each panel form a separate three-pair FDR family. Sample restriction does not establish demographic matching.**

Thresholds are displayed to three decimal places; counts use full-precision panel-specific thresholds. P-values compare continuous relative-gamma distributions, not membership proportions. FDR adjustment is within each panel; families overlap.

All tests are two-sided asymptotic Mann–Whitney tests with continuity correction. Positive rank-biserial r denotes larger values in group a (2U/(n n ) − 1). The standalone full-group and age-restricted ASD–FXS comparisons are unadjusted. All other families contain three comparisons with BH-FDR correction. A–D identify the restrictions in Figure S2. Effect estimates are descriptive and have no cluster-adjusted confidence intervals.

### Clinical assessment analyses

All clinical analyses concern the ASD cohort: 35 participants meeting the FXS-derived relative-gamma criterion (Meeting group in Table S3) and 124 below it (Below group in Table S3). Participants with FXS are not included in these analyses. Tables report all 27 included clinical measures. Bonferroni family sizes are IQ 4, SRS 7, ABC 5, SCQ Lifetime 1, and Vineland separately within assessment, subgroup, and correlation method; Welch comparisons form separate assessment-specific families. The two sensitivity restrictions also have separate families. These adjustments do not control all tests across assessments, methods, subgroups, or sensitivity sets. NE means not estimable (fewer than three paired observations or a constant variable for correlations). Raw and adjusted p-values are shown to three decimals; exact values are supplied in the companion CSV tables.

Two-sided Welch tests compare clinical scores between ASD subgroups. Mean difference is meeting minus below criterion. No comparison met the assessment-specific Bonferroni threshold. Overall IQ composite includes Leiter nonverbal IQ composites. Available IQ estimate uses a hierarchy of overall composite, abbreviated IQ, then nonverbal-domain fallback; it overlaps the overall-composite endpoint. Labels do not establish psychometric equivalence across instruments.

### Electrode mappings

Labels identify the channels used to calculate relative gamma. The GSN32 mapping began with nearest-neighbor coordinate matching and included manual Fz and Cz assignments. Channel selection followed average referencing; these mappings do not imply identical spatial sampling across net types.

---

**Table S1.** Sample-restriction results.

| Restriction | n ASD/FXS | ASD meeting criterion |
| --- | --- | --- |
| Males | 142/41 | 33/142 (23.2%) |
| Age ≥13 years | 122/41 | 32/122 (26.2%) |
| Males, age ≥13 years | 110/41 | 30/110 (27.3%) |
| Males, age ≥18 years | 64/38 | 21/64 (32.8%) |

**Table S2.** Continuous-distribution comparisons.

| Family | Comparison | n a/b | U a | Raw p | Adjusted p | Rank-biserial r |
| --- | --- | --- | --- | --- | --- | --- |
| Full groups: ASD–FXS | ASD vs FXS | 159/41 | 472 | 3.33743e-17 | 3.33743e-17 | -0.8552 |
| ASD age ≥13 years–FXS | ASD vs FXS | 122/41 | 409 | 1.25077e-15 | 1.25077e-15 | -0.8365 |
| Two-group comparison | ASD vs FXS | 159/41 | 472 | 3.33743e-17 | 1.00123e-16 | -0.8552 |
| ASD age strata–FXS | ASD <13 vs FXS | 37/41 | 63 | 3.53687e-12 | 5.3053e-12 | -0.9169 |
| ASD age strata–FXS | ASD 13–<18 vs FXS | 50/41 | 48 | 6.74427e-15 | 2.02328e-14 | -0.9532 |
| ASD age strata–FXS | ASD ≥18 vs FXS | 72/41 | 361 | 2.82945e-11 | 2.82945e-11 | -0.7554 |
| A: Males | ASD vs FXS | 142/41 | 429 | 9.94944e-17 | 2.98483e-16 | -0.8526 |
| B: Age ≥13 years | ASD vs FXS | 122/41 | 409 | 1.25077e-15 | 3.75232e-15 | -0.8365 |
| C: Males age ≥13 years | ASD vs FXS | 110/41 | 366 | 2.7605e-15 | 7.9382e-15 | -0.8377 |
| D: Males age ≥18 years | ASD vs FXS | 64/38 | 304 | 2.81204e-10 | 4.21805e-10 | -0.7500 |

**Table S3.** Clinical scores and ASD subgroup comparisons.

| Assessment | Measure | Meeting: n, mean (SD) | Below: n, mean (SD) | Mean difference | Raw p | Adjusted p |
| --- | --- | --- | --- | --- | --- | --- |
| IQ | Overall IQ composite (standard score) | 29: 83.31 (26.62) | 94: 90.88 (26.72) | -7.57 | 0.187 | 0.749 |
| IQ | Verbal-domain standard score | 18: 73.83 (22.12) | 67: 84.52 (27.75) | -10.69 | 0.095 | 0.380 |
| IQ | Nonverbal-domain standard score | 16: 77.81 (23.97) | 65: 87.91 (28.24) | -10.10 | 0.158 | 0.631 |
| IQ | Available IQ estimate (standard score; hierarchical) | 29: 83.31 (26.62) | 100: 89.83 (26.66) | -6.52 | 0.252 | 1.000 |
| SRS | SRS Total Raw | 28: 82.89 (27.80) | 88: 97.09 (25.42) | -14.20 | 0.021 | 0.145 |
| SRS | SRS Social Communication and Interaction Raw | 28: 65.71 (21.73) | 88: 78.39 (21.03) | -12.67 | 0.010 | 0.067 |
| SRS | SRS Social Awareness Raw | 29: 10.62 (3.16) | 89: 12.02 (3.72) | -1.40 | 0.052 | 0.365 |
| SRS | SRS Social Cognition Raw | 28: 15.89 (7.29) | 89: 17.19 (5.88) | -1.30 | 0.396 | 1.000 |
| SRS | SRS Social Communication Raw | 29: 27.17 (10.42) | 88: 32.59 (9.67) | -5.42 | 0.017 | 0.121 |
| SRS | SRS Social Motivation Raw | 29: 13.28 (6.35) | 89: 16.69 (5.83) | -3.41 | 0.014 | 0.098 |
| SRS | SRS Restricted/Repetitive Behavior Raw | 29: 15.83 (7.88) | 89: 18.83 (6.06) | -3.00 | 0.068 | 0.473 |
| ABC | ABC Irritability | 30: 7.23 (7.22) | 100: 9.10 (7.95) | -1.87 | 0.231 | 1.000 |
| ABC | ABC Lethargy/Social Withdrawal | 30: 11.07 (7.87) | 99: 12.35 (8.43) | -1.29 | 0.444 | 1.000 |
| ABC | ABC Stereotypy | 30: 4.80 (4.51) | 100: 4.90 (4.72) | -0.10 | 0.916 | 1.000 |
| ABC | ABC Hyperactivity | 30: 8.67 (6.33) | 101: 12.37 (8.51) | -3.70 | 0.012 | 0.061 |
| ABC | ABC Inappropriate Speech | 30: 3.40 (2.98) | 101: 3.55 (2.94) | -0.15 | 0.804 | 1.000 |
| SCQ Lifetime | SCQ Lifetime Total Score | 23: 21.87 (5.92) | 90: 20.59 (7.44) | 1.28 | 0.386 | 0.386 |
| Vineland | Vineland Adaptive Behavior Composite Standard Score | 25: 64.28 (15.42) | 84: 69.62 (15.23) | -5.34 | 0.136 | 0.543 |
| Vineland | Vineland Communication Standard Score | 25: 67.84 (18.48) | 84: 70.65 (17.39) | -2.81 | 0.502 | 1.000 |
| Vineland | Vineland Daily Living Skills Standard Score | 26: 67.54 (19.50) | 84: 74.01 (18.52) | -6.47 | 0.142 | 0.569 |
| Vineland | Vineland Socialization Standard Score | 26: 61.62 (18.01) | 84: 67.90 (17.32) | -6.29 | 0.124 | 0.497 |

**Table S4.** Net-specific labels for the 20 target electrodes.

| 10-20 target | GSN129 label | GSN32 label |
| --- | --- | --- |
| Fp1 | E22 | E1 |
| Fp2 | E9 | E2 |
| F7 | E33 | E11 |
| F3 | E24 | E3 |
| Fz | E11 | E17 |
| F4 | E124 | E4 |
| F8 | E122 | E12 |
| T7 | E45 | E13 |
| C3 | E36 | E5 |
| Cz | E55 | E28 |
| C4 | E104 | E6 |

| 10–20 target | GSN129 label | GSN32 label |
| --- | --- | --- |
| T8 | E108 | E14 |
| P7 | E58 | E15 |
| P3 | E52 | E7 |
| Pz | E62 | E19 |
| P4 | E92 | E8 |
| P8 | E96 | E16 |
| O1 | E70 | E9 |
| Oz | E75 | E20 |
| O2 | E83 | E10 |

